# Role of Combining Anticoagulant and Antiplatelet Agents in COVID-19 Treatment: A Rapid Review

**DOI:** 10.1101/2021.03.21.21253754

**Authors:** Kamal Matli, Raymond Farah, Mario Maalouf, Christy Costanian, Nibal Chamoun, Georges Ghanem

## Abstract

Although primarily affecting the respiratory system, COVID-19 causes multiple organ damage. One of its grave consequences is a prothrombotic state that manifests as thrombotic, microthrombotic, and thromboembolic events.Therefore, understanding the effect of antiplatelet and anticoagulation therapy in the context of COVID-19 treatment is important. The aim of this rapid review is to highlight the role of thrombosis in COVID-19 and provide new insights on the use of antithrombotic therapy in its management. A rapid systematic review was performed using preferred reporting items for systematic reviews. Papers published in English on antithrombotic agent use and COVID-19 complications were eligible. Results showed that the use of anticoagulants increased survival and reduced thromboembolic events in patients. However, despite the use of anticoagulants, patients still suffered thrombotic events likely due to heparin resistance. Data on antiplatelet use in combination with anticoagulants in the setting of COVID-19 is quite scarce. Current side effects of anticoagulation therapy emphasize the need to update treatment guidelines. In this rapid review, we address a possible modulatory role of antiplatelet and anticoagulant combination against COVID□19 pathogenesis. This combination may be an effective form of adjuvant therapy against COVID□19 infection. However, further studies are needed to elucidate potential risks and benefits associated with this combination.

It was not appropriate or possible to involve patients or the public in the design, or conduct, or reporting, or dissemination plans of our research

## INTRODUCTION

Coronaviruses are a major cause of public health concern due to their highly infectious nature and propensity to cause acute outbreaks as portrayed historically by severe acute respiratory syndrome (SARS) and Middle East respiratory syndrome (MERS).[1,2] Coronavirus disease 2019 (COVID-19) proves to be no exception. As of December 3^rd^ 2020, there have been more than 60 million confirmed cases and more than 1.4 million deaths worldwide related to COVID-19 according to the latest report by the World Health Organization (WHO).[3]

Although primarily affecting the respiratory system, COVID-19 causes multiple organ damage. After transmission via droplet inhalation, the virus (SARS-CoV-2) propagates within the respiratory tract and attaches to the lung epithelium via the angiotensin-converting enzyme 2 (ACE2) expressed on the cell’s surface.[4,5] Successful containment of the infection by the immune system is essential for viral eradication and disease resolution. However, some patients develop a maladaptive immune response against the virus marked by hypercytokinemia (cytokine storm), resulting in constant viral shedding and progression to advanced disease.[6] Extra-pulmonary manifestations of severe COVID-19 involve complications related to the heart, kidney, bladder, esophagus, and ileum due to elevated ACE2 expression within these organs.[7]

COVID-19 associated thrombosis and coagulopathy is a major cause of morbidity and mortality in affected patients with the underlying mechanisms incompletely understood. The disease itself causes a hypercoagulable state by altering the natural balance of circulating prothrombotic factors in severe infections. [8,9] Moreover, hyper-viscosity was demonstrated in a series of 15 critically ill patients wherein plasma viscosity as assessed by capillary viscometry was elevated.[10] Endothelial dysfunction may be the common pathophysiological process underlying these complications, as SARS-CoV-2 exhibits tropism towards the endothelial tissue and previous coronaviruses have been implicated in endothelial dysfunction as well.[8–10] Endothelial cells play a vital role in maintaining homeostasis by secreting factors that regulate blood flow, vascular tone, coagulation, and vessel wall inflammation.[11] Disruption of normal endothelial function results in an imbalance which favors a proinflammatory and procoagulant state.[12] Infection of endothelial cells by SARS-CoV-2 through ACE2 results in the internalization and downregulation of this receptor, which prevents its normal functioning. The end result of this downregulation would be an increase in prothrombotic signaling due to the accumulation of angiotensin II (Ang II).[13] SARS-CoV-2 further leads to the activation of the complement system, primarily through the lectin pathway, resulting in endothelial injury and the production of more prothrombotic factors.[12–14] It is through these diverse and interconnected systems that COVID-19 favors a prothrombotic environment which clinically manifests as thromboembolic and microthrombotic phenomena.[15]

Although further studies are required to fully understand COVID-19 etiology, coagulopathy appears to be linked to its severe manifestations and complications.[16,17] A model wherein endothelial dysfunction provides adequate explanation for COVID-19’s diverse clinical manifestations and guides future endeavors in treatment and patient management is needed.[18] Consequently, it is vital to address antiplatelet and anticoagulation therapy in this context. The aim of this rapid review is to highlight the role of thrombosis in COVID-19 and provide new insight regarding the use of antithrombotic therapy in its management.

## METHODS

This review was performed using a rapid review methodology in which the steps of a systematic review are streamlined or accelerated to produce evidence in a shortened timeframe.

A comprehensive systematic literature search was conducted to examine the possible use of anticoagulation and antiplatelet therapies in reducing thrombotic events associated with COVID-19. Four major databases were searched: OVID Medline, Web of Science, PubMed, and Google Scholar from January 2020 until November 2020. The following combination of key terms were employed: Combination 1: “COVID-19” OR “SARS” AND “platelet aggregation inhibitors” AND “anticoagulants”. Combination 2: “COVID-19” OR “SARS” AND “anticoagulants” OR “platelet aggregation inhibitors”. Moreover, bibliographies of relevant studies were hand-searched.

Studies included were in English and had clearly defined outcomes such as patient mortality, survival rates, and thrombotic status. Studies without a clearly defined untreated control group and that did not clearly define whether antithrombotic treatment was administered before or after COVID-19 infection or if it was administered as a treatment for any other comorbidity were excluded. Also, studies with a primary endpoint other than mortality and/or the development of thrombotic or thromboembolic events were excluded. Review articles, case reports, case series, commentaries, short communications, were not included. One author (RF) performed title/abstract screening and data extraction. A stringent examination of study methods was performed before data extraction to ensure the quality of each study and applicability of all eligibility criteria. Discrepancies regarding the inclusion or exclusion of articles were resolved by discussion between authors (KM, RF and CC) until consensus was reached.

A data extraction form was developed and piloted and then used to extract study details, antithrombotic therapies used, and complications and outcomes. Extracted data for each source document were verified by a second author (CC), and any discrepancies found in the verification stage were adjudicated by a third author (KM).

## RESULTS

Results from databases and bibliographic search yielded 1,577 articles **(Fig 1**.). After title and abstract screening, deduplication, and full text assessment, 12 articles met inclusion criteria (Table 1) and are presented below.

**Figure 1.**
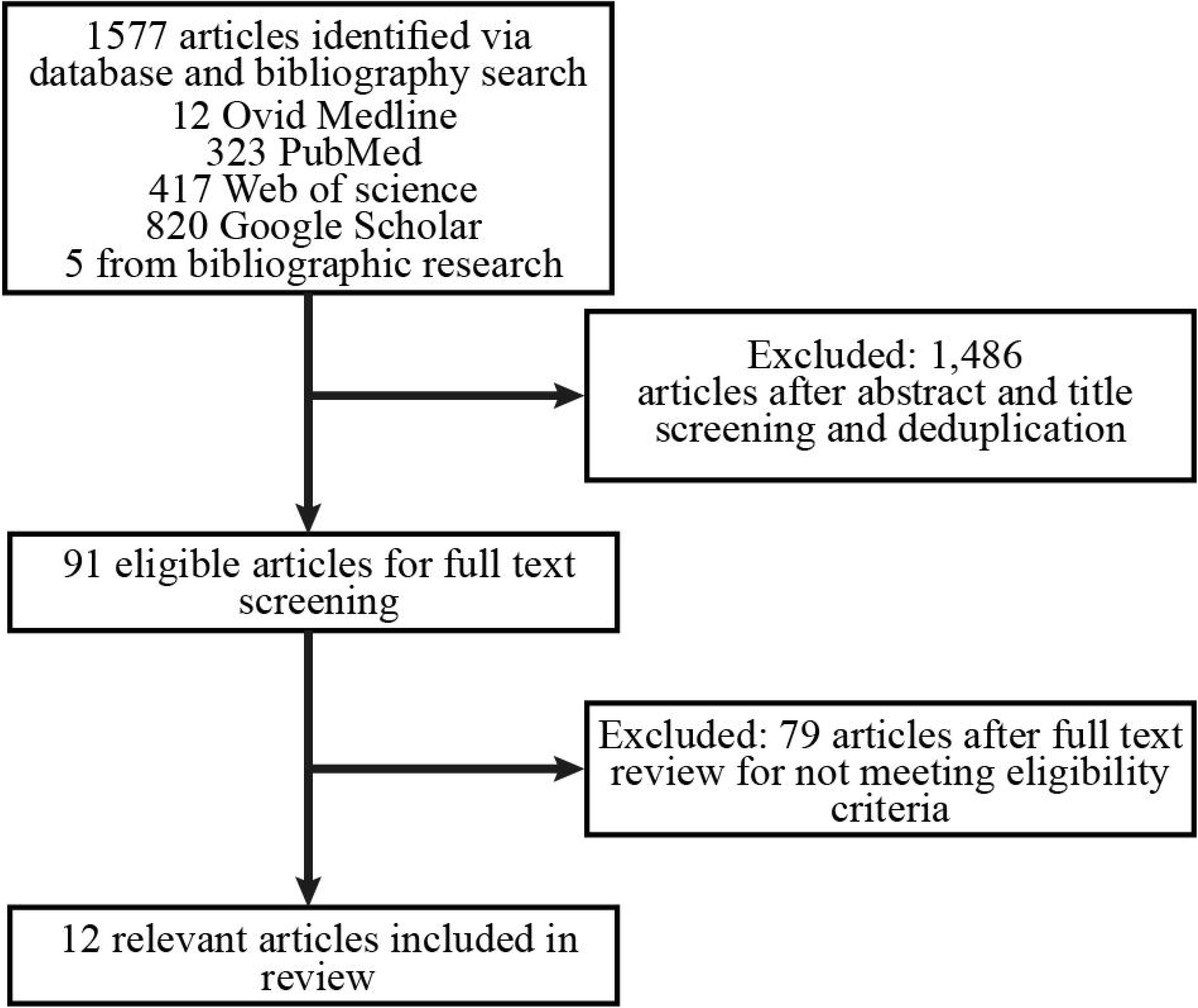
Study Flow Chart

**Table1:**
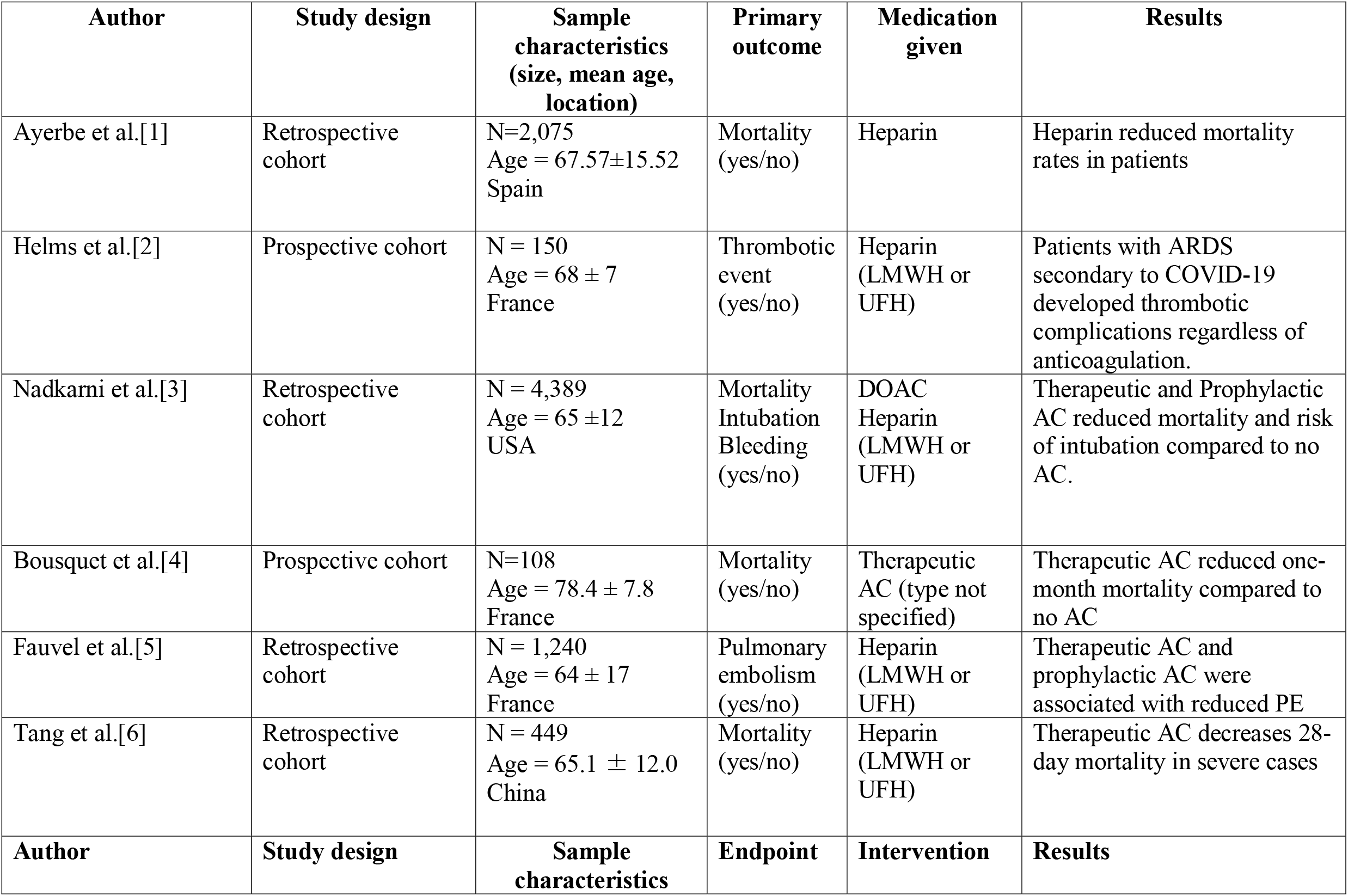
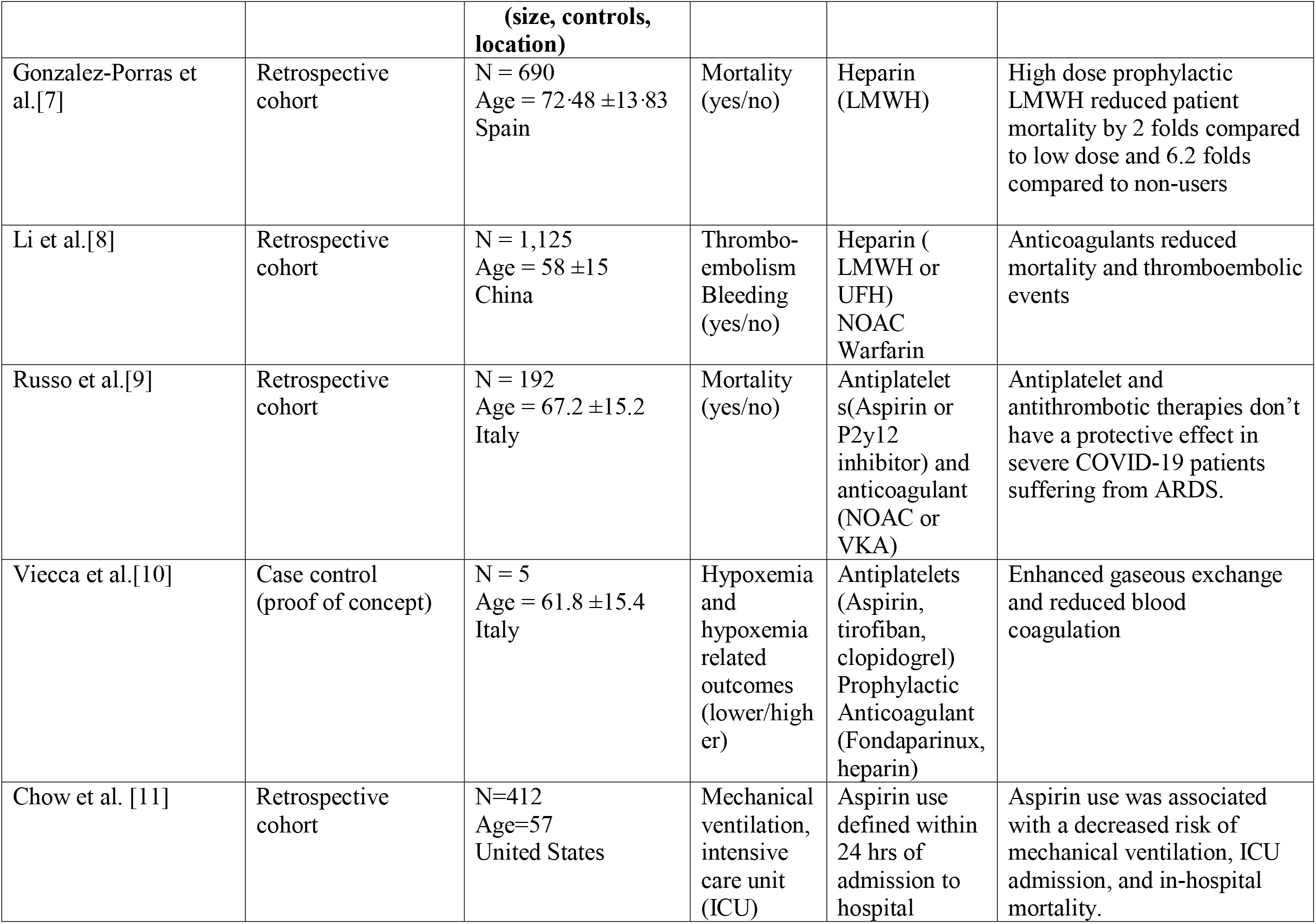
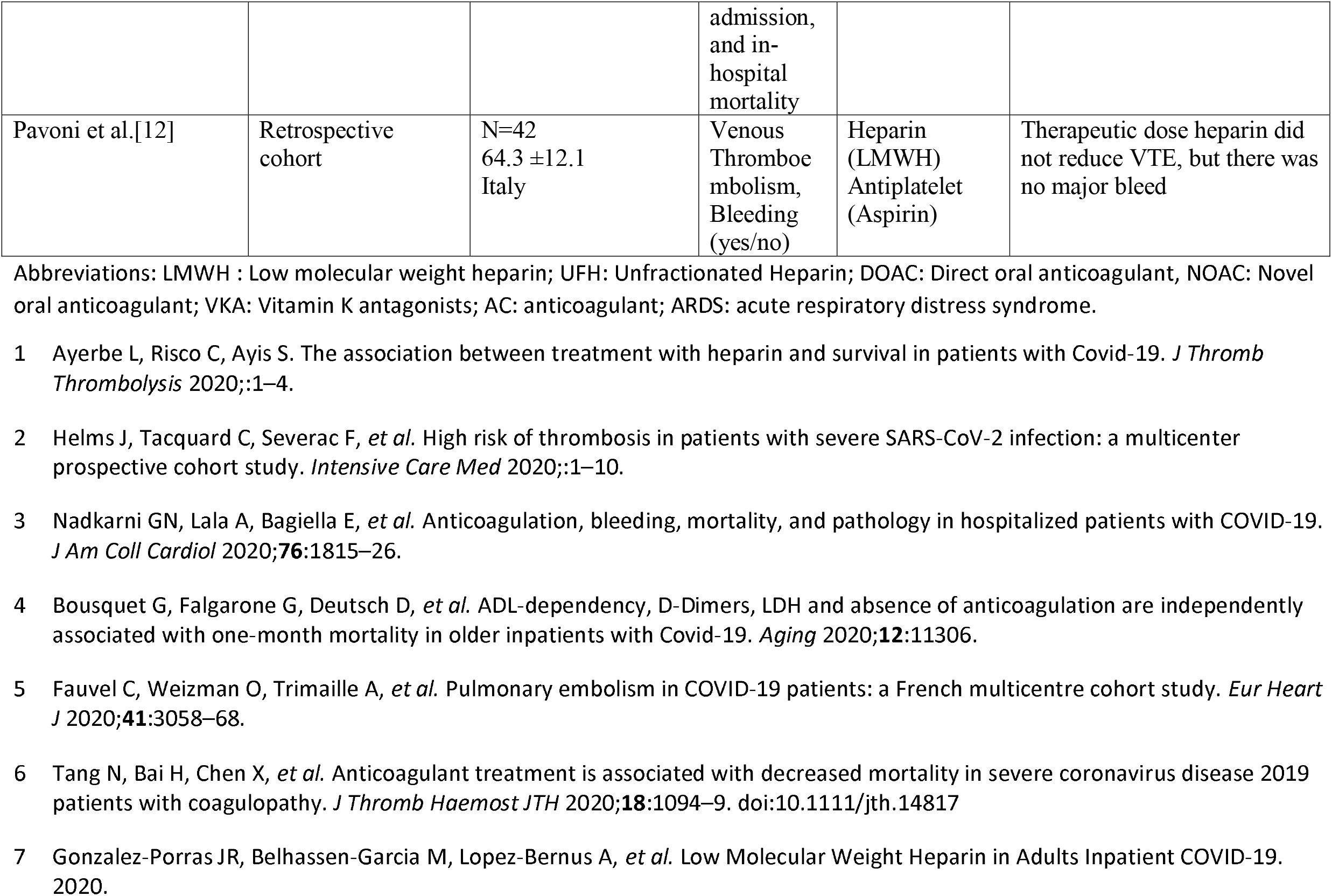
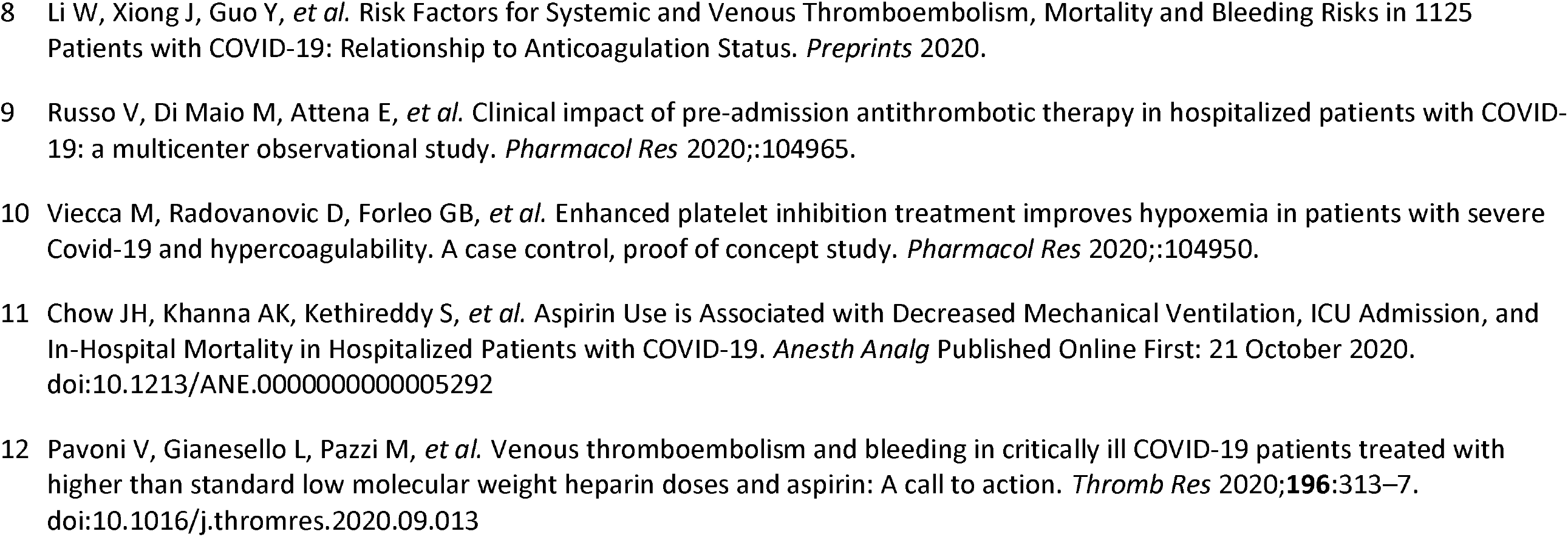
Study characteristics of papers included in this review

### Antithrombotic therapy and mortality

Nadkarni et al. investigated the impact of prophylactic and therapeutic dose heparin or direct oral anticoagulants on the mortality of 2,859 COVID-19 patients as compared to 1,530 non-anticoagulant users in USA.[19] Based on the analysis, it was evident that all patients treated with an anticoagulant had a reduced mortality when compared to those not treated. Both prophylactic and therapeutic regimens (HR: 0.53; 95% [CI]: 0.45 to 0.62, p<0.001) significantly reduced patient mortality compared to non-anticoagulant treated patients, with therapeutic dose anticoagulant (HR: 0.50; 95% CI: 0.45 to 0.57 p<0.001) being associated with a slightly reduced hazard of mortality when compared to prophylactic dose anticoagulant.[19] Interestingly, when compared to non-anticoagulant users, the incidence of intubation was reduced in both prophylactic (HR: 0.72; 95% CI: 0.58 to 0.89; p = 0.003) and therapeutic anticoagulant and (HR: 0.69; 95% CI: 0.51 to 0.94; p = 0.02) subgroups.[19] These findings were corroborated by Ayerbe et al., who found that heparin administration in 1,734 Spanish patients was associated with a statistically significant decreased risk of mortality among COVID-19 patients (age and gender adjusted OR: 0.55; 95% CI: 0.37 to\ 0.82 p = 0.003). This association remained significant even after adjustment for patient fever (>37°C) and oxygen saturation (levels < 90%) (OR: 0.54; 95% CI: 0.36 to 0.82 p = 0.003). The protective association between heparin and mortality was also independent of antiviral and adjunct treatment regimens (OR 0.42; 95% CI: 0.26 to 0.66 p < 0.001).[20] However, no data was provided whether heparin was therapeutic or prophylactic.[20] To add, by retrospectively comparing mortality of 611 patients in Spanish hospitals treated with different doses of low molecular weight heparin (LMWH) with those not receiving any anticoagulant treatment, Gonzalez-Porras et al. reported a major advantage of high dose prophylactic LMWH in reducing mortality when compared to low dose prophylactic LMWH treatment and to the complete absence of treatment.[21] Absence of anticoagulant treatment led to a 6-fold increase in patient mortality when compared to high dose LMWH (OR 6.2; 95% CI: 2.6 −14.6), and treatment with low dose LMWH was associated with a 2-fold increase in patient mortality when compared to high dose LMWH (OR 2.0; 95% CI: 1·1 to 3.6).[21] Bousquet et al. reported a 4.2-fold increase in mortality in patients not treated with anticoagulant (HR 4.20; 95% CI: 1.36-12.9), by reference to those on therapeutic doses and prophylactic treatment increases patient mortality 1.2-folds when compared to therapeutic doses (HR: 1.20; 95% CI: 0.43-3.31).[22]

On the other hand, Tang et al. reported that heparin did not reduce mortality in 99 patients receiving therapeutic anticoagulant treatment compared to non-anticoagulant group (n = 350) 30.3% vs 29.7% respectively, p = 0.910) at a Chinese hospital. This observation was seen regardless of underlying comorbidities such as diabetes, hypertension or heart disease.[23] Investigations were also done to assess the change in risk of mortality upon treatment with heparin among patients suffering from severe sepsis induced coagulopathy (SIC) and patients at high risk of developing thrombotic events given their elevated D-dimer levels (>3.0 μg/ml). Patients receiving therapeutic dose heparin exhibited a decrease in mortality when compared to their matched controls. Patients on anticoagulants having an SIC score ≥ 4 experienced a 40.0% mortality compared to 64.2% mortality in non-anticoagulant controls (p = 0.029), while patients suffering from D-dimer elevation experienced a 32.8% mortality compared to 52.4% mortality in non-AC controls (p = 0.017). [23] Similarly, Russo et al. investigated the effect of antiplatelets (acetylsalicylic acid and P2y12 inhibitor) and anticoagulants (non-vitamin K anticoagulant and vitamin K anticoagulant) on mortality of patients and on development of Acute Respiratory Distress Syndrome (ARDS) secondary to COVID-19 infection in 192 COVID-19 patients admitted into Italian hospitals.[24] They reported that regardless of antithrombotic therapy type, mortality remain unchanged. Similarly, there was no change in ARDS outcome in treated COVID-19 patients.[24] Patients in this study were not put on antiplatelet and anticoagulant combination therapy; therefore, the synergistic effect of these therapies could not be analysed.[24]

### Antithrombotic therapy and thrombotic events

Thromboembolic events namely, deep vein thrombosis (DVT) and pulmonary embolisms (PE), are involved in COVID-19 pathology. Available data indicates presence of an association between thromboembolic events and higher patient mortality.[25] Li et al. investigated the possible therapeutic value of anticoagulants (Heparin, Novel oral anticoagulant, NOAC, and Warfarin) administered orally or parenterally[26] among 1,125 patients admitted to a Chinese hospital. Results showed that the administration of oral or parenteral anticoagulant successfully reduced the risk of venous thrombosis in 249 treated patients when compared to those not treated (p < 0.01). Moreover, only heparin significantly reduced the risk of thromboembolism, bleeding and mortality as a composite outcome (HR: 0.70; 95% CI: 0.51 to 0.95, p=0.02). However, heparin use was not associated with mortality, but was associated with a borderline reduction in thromboembolism and bleeding (HR: 0.36; 95% CI: 0.13 to 1.01, p=0.053). These observed effects were independent of COVID-19 infection severity, presence of comorbidities, surgery, antiviral drugs, immunomodulators, Chinese herb use and antiplatelet medication (Aspirin and Clopidogrel).[26] Fauvel et al. described the benefit of administering prophylactic or therapeutic doses of heparin in reducing the incidence of pulmonary embolism (PE) by assessing 1,240 COVID-19 patients admitted to multiple French hospitals. Therapeutic dose heparin was associated with a decreased risk of PE (OR: 0.87; 95% CI 0.82 to 0.92, P < 0.001) and prophylactic dose was also associated with a decreased risk of PE (OR: 0.83; 95% CI: 0.79 to 0.85, P < 0.001).[27] It was evident that the incidence of PE in COVID-19 patients was also associated with higher levels of C-reactive protein (OR: 1.03; 95% CI: 1.01–1.04; p = 0.001).[27]

Helms et al. investigated the possible benefit of therapeutic and prophylactic dose heparin in 150 COVID-19 patients in two French hospitals and suffering from ARDS and presenting with elevated fibrinogen levels and systemic inflammation.[28] These patients were matched to non-COVID-19 ARDS patients to compare the occurrence of thromboembolic complications and PE in the two groups. Contrary to previous findings, it was evident that despite prophylactic or therapeutic anticoagulation with heparin, patients suffering from COVID-19 ARDS were more likely to develop thromboembolic complications (OR: 2.6; 95% [CI]: 1.1 to 6.1, p = 0.04) and pulmonary embolisms (OR : 6.2; 95% [CI]: 1.6–23.4; p = 0.01) when compared to their non-COVID-19 ARDS controls.[28] Moreover, D-dimer levels were higher in COVID-19 ARDS patients by a mean 4.5-fold increase, than in non-COVID-19 patients (p < 0.001).[28]

### Antiplatelet use

The use of antiplatelets has also shown benefit among COVID-19 patients, especially in alleviating respiratory symptoms.[29] In a case-control study conducted in Italy, five patients presenting with more than a 3-fold increase in D-dimer levels, were placed on fondaparinux and an antiplatelet therapy regimen (Aspirin and/or clopidogrel and a continuous infusion of tirofiban), while controls received standard prophylactic heparin infusions only. Treatment with antiplatelets progressively decreased Alveolar-arterial O2 gradient by 138 (49) mmHg (p = 0.005) and progressively increased PAO2/FIO2 by 108 (57) mmHg (p = 0.037) during the 7-day observation period. To add, C-reactive protein levels were reduced from 62 mg/dL at baseline to 28 mg/dL in response to treatment after 7 days (p = 0.044), thus indicating a possible role of combined antiplatelet therapy in reducing endothelial inflammation.[29] However, the small sample size of this study does not allow drawing of valid inferences and more studies with higher power are warranted. Moreover, a retrospective cohort study conducted by Chow et al[30] revealed that patients who had received aspirin within 24 hours of hospital admission had decreased rates of mechanical ventilation (adjusted HR: 0.56; 95% [CI]: 0.37-0.85; p=0.007), ICU admission (adjusted HR: 0.57; 95% [CI]: 0.38-0.85; p=0.005), and in-hospital mortality (adjusted HR 0.53; 95% [CI]: 0.31-0.90; p=0.02). No significant difference was observed between patients who received aspirin and those who did not, in terms of major bleeding (p=0.69) or overt thrombosis (p=0.82).[30] In addition, a retrospective study among 42 patients admitted to the ICU with COVID-19, and who received full dose or prophylactic dose of anticoagulation along with aspirin reported no increase in major bleeds.[31]

## DISCUSSION

Amid the COVID-19 pandemic, consistent treatment guidelines for physicians remain farfetched. An important aspect of COVID-19 progression is the development of coagulopathy which necessitates the adoption of proper antithrombosis treatment plans that could be administered to patients suffering from mild, moderate to severe disease. With current guidelines emphasizing the importance of thromboprophylaxis in both high and low risk COVID-19 patients, it has become imperative to assess the efficacy and proper prescription of such treatments.[32–34] Overall, the use of anticoagulants has shown an increased survival and reduction in thromboembolic events in patients receiving treatment when compared to untreated controls. Heparin, whether unfractionated heparin (UFH) or LMWH, was the most common choice of anticoagulant for management of COVID-19 associated thrombotic events. On the whole, combined antiplatelet and anticoagulant therapy increased patient survival and alleviated respiratory symptoms secondary to PE, which indicates anq advantage for combination therapy over antiplatelet or anticoagulant therapy alone[24].

### Prophylactic vs. Therapeutic Anticoagulants

Clinically, when prescribing anticoagulant regimens, physicians attempt to achieve a balance between healthy blood coagulation and bleeding risk. This balance is contested in COVID-19 as multiple patients develop a hypercoagulable state despite anticoagulation use. The available data indicates that anticoagulation presents patients with a clear advantage over absence of anticoagulation, regardless of minor bleeding risk that accompanies the treatment.[19–23,26,27] Studies assessing the efficacy of therapeutic anticoagulation over prophylactic are still rudimentary, but data seems to indicate that therapeutic doses reduce thromboembolic events and reduce mortality risk when compared to prophylactic doses.[19,35–37] Therapeutic dose enoxaparin significantly increased PO2/FiO2 ratio, (PO2/FiO2 = 261 [95% CI 230–293], p=0.0004) when compared to patients placed on prophylactic anticoagulation. Ultimately, these patients were also weaned off of ventilation (HR : 4.0 ; 95% [CI]: 1.035 to 15.053], p = 0.031).[36] Moreover, therapeutic dose anticoagulation seems to reduce endothelial cell lesion (p = 0.02) which could also reduce the thromboembolic risk of COVID-19, suggesting another therapeutic target for anticoagulants.[37]

### Bleeding events

Current evidence on the risk of increased bleeding events in COVID-19 patients receiving therapeutic or prophylactic anticoagulant treatment remains inconclusive. In this context, some data suggest that bleeding events increase in patients placed on antithrombotic therapy especially when comparing high dose therapies to lower doses (p < 0.003).[19,21,38] However, other studies indicated lower bleeding incidents in patients treated with therapeutic or prophylactic dose anticoagulant when compared to non-users.[23,26] This necessitates further assessment to ascertain whether bleeding risk due to anticoagulant use outweighs the benefits.

### C-reactive protein

Knowing that C-reactive proteins play an important role in activating the blood complement system, and that the latter in turn can mediate coagulation, increased C-reactive protein levels reported in COVID-19 patients might reflect increased levels of systemic coagulopathy, along with increased inflammatory responses that could be attributed to endothelial dysfunction.[19,27,39,40] Complement activation has been well reported in COVID-19 patients, but its root causes remain to be properly delineated.[41–43]

### Heparin resistance

A possible explanation for the failure of heparin to properly inhibit or reduce coagulation in COVID-19 patients could be the development of heparin resistance in a select group of patients suffering from aggravated disease status. Patients prone to heparin resistance commonly present with a deficiency in antithrombin III, increased fibrinogen and D-dimer level.[44,45] Both fibrinogen levels and D-dimer levels have been shown to be elevated in COVID-19 patients, especially in severe cases who develop thromboembolic complications, and who only respond to higher doses of antithrombotic treatment.[9,21– 23,28,29,46,47] Additionally, heparin resistance has been reported in COVID-19 patients.[48,49] and could therefore explain the failure of anti-thrombotic therapies in some patients in reducing coagulopathy. The administration of antithrombin III proved useful in abolishing heparin resistance in patients who underwent cardiac surgery and could therefore possibly benefit COVID-19 patients.[45] The impact of heparin resistance should be assessed in COVID-19 patients to identify whether it only affects a few isolated cases or it is actually a key player in the COVID-19 induced coagulopathy.

### D- dimers

When a blood clot is dissolved, D-dimers are disseminated in the blood as a biproduct of coagulated platelet break-down.[50] Given that coagulopathy is not uncommon in COVID-19 patients, it is expected that D-dimer levels might play an important role in diagnosis or monitoring of patients. Current studies observed elevated D-dimer levels in COVID-19 patients, ranging from a 2.5-fold to a 6-fold increase, with these increased levels predicting mortality and the development of thromboembolic events in patients.[21–23,28,29,46] Yet, there is no consensus on a specific cut-off level which can accurately predict disease course, mortality or response to antithrombotic therapy. Mouhat et al. suggested the use of 2,590 ng/mL as threshold level which corresponds to ≈ 5-fold increase in normal D-dimer level.[51] This 5-fold increase was associated with a 17-fold increase in the incidence of pulmonary embolism among patients (OR: 16.9, 95% [CI]: 6.3 to 45.0, p < 0.001).[51] Similarly, Ventura-Diaz et al., reported that a 5.5-fold increase in D-dimer level could predict the occurrence of PE with 81% sensitivity, and 59% specificity (p < 0.001),[52] while, Ooi et al. examined a larger sample of 974 patients and reported that a 4.5-fold increase in D-dimer level would predict the occurrence of PE with 72% sensitivity and a 74% specificity.[46] Moreover, patients presenting with increased D-dimer levels equivalent to more than 4-folds the normal level exhibit a higher mortality risk when compared to their counterparts who present with lower levels (OR: 10.17; 95% [CI]: 1.10–94.38, P = 0.041].[53] These findings were corroborated by Zhang et al. who had similar findings and showed that D-dimer levels ≥ 4 times the normal level could predict mortality with 92.3% sensitivity 83.3%. specificity (HR: 51.5; 95% [CI]: 12.9 to 206.7, p< 0.001).[54] further research on possible explanations for anticoagulant treatment failure in some patient subgroups is warranted.[23] Collectively, these preliminary findings indicate the possible use of D-dimers to guide treatment regimens and to establish different patient subgroups who should receive personalized treatment. However, the use of D-dimer in clinical settings as a stand-alone marker for COVID-19 thrombosis does not seem practical, as a clear cut-off value cannot be easily established since sensitivity of the test is compromised with age.[55,56]

### Arterial thrombotic events

Apart from the venous thrombotic and thromboembolic events, there are also reports of arterial thrombosis including stroke, myocardial infarction, acute limb ischemia, aortic thrombosis, and splenic infarcts.[57–60] Microvascular thrombosis also is present in COVID-19 disease. Autopsies done on patients who died from COVID-19 infection demonstrated microvascular thrombosis in the lungs. The mechanism of development of this entity is unclear and is thought to be multifactorial.[61–64]

### Commentary

Coagulation is a highly well-organized procedure that involves the interaction of endothelial cells, platelets, and coagulation factors.[65] Under physiologic conditions platelets circulate without adhering to intact and inactive endothelium. COVID-19 infection was shown to be highly associated with endothelial dysfunction.[66–68] Thus, when endothelial activation and dysfunction occur, disruption of vascular integrity and endothelial cell apoptosis results in exposure of the thrombogenic basement membrane and activation of the clotting cascade by displaying vWF, P-selectin, and fibrinogen, onto which activated platelets bind and play their primary role in thrombosis.[21,69,70] Those activated platelets also produce VEGF, which induces endothelial cells to express the tissue factor, the main activator of the coagulation cascade. The coagulation pathway is also activated when the blood vessels are injured. The transfer of microthrombi into the systemic circulation increases the risk of development of deep vein thrombosis.[71] Even though the underlying mechanisms of thrombosis in COVID-19 is incompletely understood, the major contributors of damage are caused by endothelial injury and hypercoagulability.[72,73] So far, anticoagulation was associated with better outcomes in COVID-19 patients with many societies recommending its use as part of the treatment in most COVID patients.[19–21] However these results were not consistent among all studies, with some reporting no added value[23,24] and others the occurrence of thrombotic events on anticoagulation.[48,49] Recently, increased interest in the use antiplatelets has surfaced for treating and preventing thrombotic events wherein there has been some benefit associated with aspirin use.[30]The dual effect of antiplatelet and anticoagulant therapy on COVID-19 induced platelet thrombosis and hypercoagulability, respectively, may result in a synergistic and superior outcome than the use of either medication alone. Especially considering that thrombotic manifestations of COVID-19 are heterogenous since they arise from several pathological mechanisms occurring simultaneously. Using a drug combination is not unheard of as it has already been studied in acute coronary syndrome (ACS) patients.[74]

## CONCLUSION

Current guidelines recommend administration of anticoagulants with varying doses based on case severity in COVID-19 patients.[75,76] However, the evidence in support of these guidelines is limited due to the paucity of high-quality clinical trials performed to date. Despite the fact that COVID-19 patients treated with anticoagulants had overall lower risk of thrombotic events and mortality, the use of anticoagulants or antiplatelets alone was still associated with the occurrence of thrombotic and microthrombotic events in a substantial number of patients. Given the aforementioned shortcomings of anticoagulation when used alone and results from this rapid review showing that combined anticoagulant and antiplatelet therapy in the treatment of COVID-19 despite being poorly studied, implied a better clinical outcome when compared with the use of anticoagulation alone without the occurrence of major bleeding. So, we recommend further clinical trials to evaluate the safety and efficacy of combining antiplatelet and anticoagulants agents in the management of COVID-19 patients.

## Data Availability

none

## Author Contributions

KM contributed to hypothesis conception and wrote final draft of manuscript. RF contributed to searching, data extraction, wrote manuscript text and managed references. MM wrote manuscript text and managed references. CC supervised searching, data extraction, and manuscript writing and editing. GG and NC contributed to critical revisions of the manuscript. All authors read and approved the final version of the manuscript before submission for publication.

## References

1 Hu B, Guo H, Zhou P, et al. Characteristics of SARS-CoV-2 and COVID-19. Nat Rev Microbiol 2020;:1–14. doi:10.1038/s41579-020-00459-7

2 Lessons of Past Coronavirus Pandemics. Popul Dev Rev 2020;46:633–7. doi:10.1111/padr.12360

3 WHO Coronavirus Disease (COVID-19) Dashboard. https://covid19.who.int (accessed 3 Dec 2020).

4 Yuki K, Fujiogi M, Koutsogiannaki S. COVID-19 pathophysiology: A review. Clin Immunol Orlando Fla 2020;215:108427. doi:10.1016/j.clim.2020.108427

5 Mason RJ. Pathogenesis of COVID-19 from a cell biology perspective. Eur Respir J 2020;55. doi:10.1183/13993003.00607-2020

6 Perico L, Benigni A, Casiraghi F, et al. Immunity, endothelial injury and complement-induced coagulopathy in COVID-19. Nat Rev Nephrol 2020;:1–19. doi:10.1038/s41581-020-00357-4

7 Zou X, Chen K, Zou J, et al. Single-cell RNA-seq data analysis on the receptor ACE2 expression reveals the potential risk of different human organs vulnerable to 2019-nCoV infection. Front Med 2020;14:185–92. doi:10.1007/s11684-020-0754-0

8 Panigada M, Bottino N, Tagliabue P, et al. Hypercoagulability of COVID-19 patients in intensive care unit: A report of thromboelastography findings and other parameters of hemostasis. J Thromb Haemost JTH 2020;18:1738–42. doi:10.1111/jth.14850

9 Ranucci M, Ballotta A, Di Dedda U, et al. The procoagulant pattern of patients with COVID-19 acute respiratory distress syndrome. J Thromb Haemost 2020. doi: 10.1111/jth.14854.

10 Maier CL, Truong AD, Auld SC, et al. COVID-19-associated hyperviscosity: a link between inflammation and thrombophilia? Lancet Lond Engl 2020;395:1758–9. doi:10.1016/S0140-6736(20)31209-5

11 Pearson JD. Normal endothelial cell function. Lupus 2000;9:183–8. doi:10.1191/096120300678828299

12 Biswas I, Khan GA. Endothelial Dysfunction in Cardiovascular Diseases. Basic Clin Underst Microcirc Published Online First: 4 November 2019. doi:10.5772/intechopen.89365

13 Miesbach W. Pathological Role of Angiotensin II in Severe COVID-19. TH Open Companion J Thromb Haemost 2020;4:e138.#x2013;44. doi:10.1055/s-0040-1713678

14 Java A, Apicelli AJ, Liszewski MK, et al. The complement system in COVID-19: friend and foe? JCI Insight 2020;5. doi:10.1172/jci.insight.140711

15 Coccheri S. COVID-19: The crucial role of blood coagulation and fibrinolysis. Intern Emerg Med 2020;15:1369–73. doi:10.1007/s11739-020-02443-8

16 Mondal S, Quintili AL, Karamchandani K, et al. Thromboembolic disease in COVID-19 patients: A brief narrative review. J Intensive Care 2020;8:70. doi:10.1186/s40560-020-00483-y

17 Ra C, Wh F. Thrombotic Complications of COVID-19 Infection: A Review. Cardiol. Rev. 2021;29. doi:10.1097/CRD.0000000000000347

18 Libby P, Lüscher T. COVID-19 is, in the end, an endothelial disease. Eur Heart J 2020;41:3038–44. doi:10.1093/eurheartj/ehaa623

19 Nadkarni GN, Lala A, Bagiella E, et al. Anticoagulation, bleeding, mortality, and pathology in hospitalized patients with COVID-19. J Am Coll Cardiol 2020;76:1815–26. doi: 10.1016/j.jacc.2020.08.041

20 Ayerbe L, Risco C, Ayis S. The association between treatment with heparin and survival in patients with Covid-19. J Thromb Thrombolysis 2020;:1–4. doi: 10.1007/s11239-020-02162-z

21 Gonzalez-Porras JR, Belhassen-Garcia M, Lopez-Bernus A, et al. Low Molecular Weight Heparin in Adults Inpatient COVID-19. 2020. doi: 10.1016/j.jvsv.2020.06.006

22 Bousquet G, Falgarone G, Deutsch D, et al. ADL-dependency, D-Dimers, LDH and absence of anticoagulation are independently associated with one-month mortality in older inpatients with Covid-19. Aging 2020;12:11306. doi: 10.18632/aging.103583

23 Tang N, Bai H, Chen X, et al. Anticoagulant treatment is associated with decreased mortality in severe coronavirus disease 2019 patients with coagulopathy. J Thromb Haemost 2020;18:1094–9. doi: 10.1111/jth.14817

24 Russo V, Di Maio M, Attena E, et al. Clinical impact of pre-admission antithrombotic therapy in hospitalized patients with COVID-19: a multicenter observational study. Pharmacol Res 2020;:104965. doi: 10.1016/j.phrs.2020.104965

25 Malas MB, Naazie IN, Elsayed N, et al. Thromboembolism risk of COVID-19 is high and associated with a higher risk of mortality: A systematic review and meta-analysis. EClinicalMedicine 2020;29:100639. doi: 10.1016/j.eclinm.2020.100639

26 Li W, Xiong J, Guo Y, et al. Risk Factors for Systemic and Venous Thromboembolism, Mortality and Bleeding Risks in 1125 Patients with COVID-19: Relationship to Anticoagulation Status. Preprints 2020.

27 Fauvel C, Weizman O, Trimaille A, et al. Pulmonary embolism in COVID-19 patients: a French multicentre cohort study. Eur Heart J 2020;41:3058–68. doi: 10.1093/eurheartj/ehaa500.

28 Helms J, Tacquard C, Severac F, et al. High risk of thrombosis in patients with severe SARS-CoV-2 infection: a multicenter prospective cohort study. Intensive Care Med 2020;:1–10. doi: 10.1007/s00134-020-06062-x

29 Viecca M, Radovanovic D, Forleo GB, et al. Enhanced platelet inhibition treatment improves hypoxemia in patients with severe Covid-19 and hypercoagulability. A case control, proof of concept study. Pharmacol Res 2020;:104950. doi: 10.1016/j.phrs.2020.104950.

30 Chow JH, Khanna AK, Kethireddy S, et al. Aspirin Use is Associated with Decreased Mechanical Ventilation, ICU Admission, and In-Hospital Mortality in Hospitalized Patients with COVID-19. Anesth Analg Published Online First: 21 October 2020. doi:10.1213/ANE.0000000000005292

31 Pavoni V, Gianesello L, Pazzi M, et al. Venous thromboembolism and bleeding in critically ill COVID-19 patients treated with higher than standard low molecular weight heparin doses and aspirin: A call to action. Thromb Res 2020;196:313–7. doi:10.1016/j.thromres.2020.09.013

32 Thachil J, Tang N, Gando S, et al. ISTH interim guidance on recognition and management of coagulopathy in COVID-19. J Thromb Haemost 2020;18:1023–6. doi:10.1111/jth.14810.

33 Moores LK, Tritschler T, Brosnahan S, et al. Prevention, diagnosis and treatment of venous thromboembolism in patients with COVID-19: CHEST Guideline and Expert Panel Report. Chest 2020. doi:10.1016/j.chest.2020.05.559

34 Casini A, Alberio L, Angelillo-Scherrer A, et al. Suggestions for thromboprophylaxis and laboratory monitoring for in-hospital patients with COVID-19. Swiss Med Wkly 2020;150:w20247. doi: 10.4414/smw.2020.20247

35 Maatman TK, Jalali F, Feizpour C, et al. Routine venous thromboembolism prophylaxis may be inadequate in the hypercoagulable state of severe coronavirus disease 2019. Crit Care Med 2020. doi:10.1097/CCM.0000000000004466

36 Lemos ACB, do Espírito Santo Da, Salvetti MC, et al. Therapeutic versus prophylactic anticoagulation for severe COVID-19: A randomized phase II clinical trial (HESACOVID). Thromb Res 2020;196:359–66. doi: https://doi.org/10.1016/j.thromres.2020.09.026

37 Khider L, Gendron N, Goudot G, et al. Curative anticoagulation prevents endothelial lesion in COVID-19 patients. J Thromb Haemost 2020;18:2391–9. doi: 10.1111/jth.14968.

38 Musoke N, Lo KB, Albano J, et al. Anticoagulation and bleeding risk in patients with COVID-19. Thromb Res 2020;196:227–30. doi: 10.1016/j.thromres.2020.08.035

39 Kenawy HI, Boral I, Bevington A. Complement-Coagulation Cross-Talk: A Potential Mediator of the Physiological Activation of Complement by Low pH. Front Immunol 2015;6. doi:10.3389/fimmu.2015.00215

40 Xu P, Lin S, Yang X, et al. C-reactive protein enhances activation of coagulation system and inflammatory response through dissociating into monomeric form in antineutrophil cytoplasmic antibody-associated vasculitis. BMC Immunol 2015;16:10–10. doi:10.1186/s12865-015-0077-0

41 Showers CR, Nuovo GJ, Lakhanpal A, et al. A Covid-19 Patient with Complement-Mediated Coagulopathy and Severe Thrombosis. Pathobiology Published Online First: 2020. doi:10.1159/000512503

42 Holter JC, Pischke SE, de Boer E, et al. Systemic complement activation is associated with respiratory failure in COVID-19 hospitalized patients. Proc Natl Acad Sci U A 2020;117:25018–25. doi:10.1073/pnas.2010540117

43 Mastellos DC, Pires da Silva Bgp, Fonseca BAL, et al. Complement C3 vs C5 inhibition in severe COVID-19: Early clinical findings reveal differential biological efficacy. Clin Immunol 2020;220:108598. doi:10.1016/j.clim.2020.108598

44 Durrani J, Malik F, Ali N, et al. To be or not to be a case of heparin resistance. J Community Hosp Intern Med Perspect 2018;8:145–8. doi:10.1080/20009666.2018.1466599

45 Kawatsu S, Sasaki K, Sakatsume K, et al. Predictors of heparin resistance before cardiovascular operations in adults. Ann Thorac Surg 2018;105:1316–21. doi: 10.1016/j.athoracsur.2018.01.068

46 Ooi M, Rajai A, Patel R, et al. Pulmonary thromboembolic disease in COVID-19 patients on CT pulmonary angiography–Prevalence, pattern of disease and relationship to D-dimer. Eur J Radiol 2020;132:109336. doi: 10.1016/j.ejrad.2020.109336

47 Iba T, Levy JH, Connors JM, et al. The unique characteristics of COVID-19 coagulopathy. Crit Care 2020;24:360. doi:10.1186/s13054-020-03077-0

48 Baccellieri D, Bilman V, Apruzzi L, et al. A case of Covid-19 patient with acute limb ischemia and heparin resistance. Ann Vasc Surg 2020;68:88–92. doi: 10.1016/j.avsg.2020.06.046

49 White D, MacDonald S, Bull T, et al. Heparin resistance in COVID-19 patients in the intensive care unit. J Thromb Thrombolysis 2020;:1. doi: 10.1007/s11239-020-02145-0

50 Bounds EJ, Kok SJ. D Dimer. In: StatPearls [Internet]. StatPearls Publishing 2019.

51 Mouhat B, Besutti M, Bouiller K, et al. Elevated D-dimers and lack of anticoagulation predict PE in severe COVID-19 patients. Eur Respir J 2020;56. doi:10.1183/13993003.01811-2020

52 Ventura-Díaz S, Quintana-Pérez JV, Gil-Boronat A, et al. A higher D-dimer threshold for predicting pulmonary embolism in patients with COVID-19: a retrospective study. Emerg Radiol 2020;:1–11. doi: 10.1007/s10140-020-01859-1

53 Yao Y, Cao J, Wang Q, et al. D-dimer as a biomarker for disease severity and mortality in COVID-19 patients: a case control study. J Intensive Care 2020;8:1–11. doi: 10.1186/s40560-020-00466-z

54 Zhang L, Yan X, Fan Q, et al. D-dimer levels on admission to predict in-hospital mortality in patients with Covid-19. J Thromb Haemost 2020;18:1324–9. doi: 10.1111/jth.14859

55 Righini M, Van Es J, Den Exter PL, et al. Age-adjusted D-dimer cutoff levels to rule out pulmonary embolism: the ADJUST-PE study. Jama 2014;311:1117–24. doi:10.1001/jama.2014.2135

56 Verma N, Willeke P, Bicsán P, et al. [Age-adjusted D-dimer cut-offs to diagnose thromboembolic events: validation in an emergency department]. Med Klin Intensivmed Notfallmedizin 2014;109:121–8. doi:10.1007/s00063-013-0265-8

57 Bilaloglu S, Aphinyanaphongs Y, Jones S, et al. Thrombosis in Hospitalized Patients With COVID-19 in a New York City Health System. JAMA 2020;324:799–801. doi:10.1001/jama.2020.13372

58 Qureshi AI, Abd-Allah F, Al-Senani F, et al. Management of acute ischemic stroke in patients with COVID-19 infection: Insights from an international panel. Am J Emerg Med 2020;38:1548.e5-1548.e7. doi:10.1016/j.ajem.2020.05.018

59 Bellosta R, Luzzani L, Natalini G, et al. Acute limb ischemia in patients with COVID-19 pneumonia. J Vasc Surg 2020;72:1864. doi:10.1016/j.jvs.2020.04.483

60 de Roquetaillade C, Chousterman BG, Tomasoni D, et al. Unusual arterial thrombotic events in Covid-19 patients. Int J Cardiol 2021;323:281–4. doi:10.1016/j.ijcard.2020.08.103

61 Magro C, Mulvey JJ, Berlin D, et al. Complement associated microvascular injury and thrombosis in the pathogenesis of severe COVID-19 infection: A report of five cases. Transl Res J Lab Clin Med 2020;220:1–13. doi:10.1016/j.trsl.2020.04.007

62 Connors JM, Levy JH. Thromboinflammation and the hypercoagulability of COVID-19. J Thromb Haemost JTH 2020;18:1559–61. doi:10.1111/jth.14849

63 Menter T, Haslbauer JD, Nienhold R, et al. Postmortem examination of COVID-19 patients reveals diffuse alveolar damage with severe capillary congestion and variegated findings in lungs and other organs suggesting vascular dysfunction. Histopathology 2020;77:198–209. doi:10.1111/his.14134

64 Ackermann M, Verleden SE, Kuehnel M, et al. Pulmonary Vascular Endothelialitis, Thrombosis, and Angiogenesis in Covid-19. N Engl J Med 2020;383:120–8. doi:10.1056/NEJMoa2015432

65 Transfusion Medicine and Hemostasis −3rd Edition. https://www.elsevier.com/books/transfusion-medicine-and-hemostasis/shaz/978-0-12-813726-0 (accessed 20 Jan 2021).

66 Gavriilaki E, Anyfanti P, Gavriilaki M, et al. Endothelial Dysfunction in COVID-19: Lessons Learned from Coronaviruses. Curr Hypertens Rep 2020;22. doi:10.1007/s11906-020-01078-6

67 Nägele MP, Haubner B, Tanner FC, et al. Endothelial dysfunction in COVID-19: Current findings and therapeutic implications. Atherosclerosis 2020;314:58–62. doi:10.1016/j.atherosclerosis.2020.10.014

68 Huertas A, Montani D, Savale L, et al. Endothelial cell dysfunction: a major player in SARS-CoV-2 infection (COVID-19)? Eur Respir J 2020;56. doi:10.1183/13993003.01634-2020

69 Sturtzel C. Endothelial Cells. Adv Exp Med Biol 2017;1003:71–91. doi:10.1007/978-3-319-57613-8_4

70 Nachman RL, Rafii S. Platelets, petechiae, and preservation of the vascular wall. N Engl J Med 2008;359:1261–70. doi:10.1056/NEJMra0800887

71 Jin Y, Ji W, Yang H, et al. Endothelial activation and dysfunction in COVID-19: from basic mechanisms to potential therapeutic approaches. Signal Transduct Target Ther 2020;5:293. doi:10.1038/s41392-020-00454-7

72 Esmon CT. Basic mechanisms and pathogenesis of venous thrombosis. Blood Rev 2009;23:225–9. doi:10.1016/j.blre.2009.07.002

73 Wolberg AS, Rosendaal FR, Weitz JI, et al. Venous thrombosis. Nat Rev Dis Primer 2015;1:15006. doi:10.1038/nrdp.2015.6

74 Eikelboom JW, Anand SS, Malmberg K, et al. Unfractionated heparin and low-molecular-weight heparin in acute coronary syndrome without ST elevation: a meta-analysis. Lancet Lond Engl 2000;355:1936–42. doi:10.1016/S0140-6736(00)02324-2

75 Marietta M, Ageno W, Artoni A, et al. COVID-19 and haemostasis: a position paper from Italian Society on Thrombosis and Haemostasis (SISET). Blood Transfus Trasfus Sangue 2020;18:167–9. doi:10.2450/2020.0083-20

76 Spyropoulos AC, Levy JH, Ageno W, et al. Scientific and Standardization Committee communication: Clinical guidance on the diagnosis, prevention, and treatment of venous thromboembolism in hospitalized patients with COVID-19. J Thromb Haemost JTH 2020;18:1859–65. doi:10.1111/jth.14929

